# Convolutional neural networks for quantitative smartphone video nystagmography: ConVNG

**DOI:** 10.1101/2022.07.24.22277934

**Authors:** M. Friedrich, E. Schneider, M. Buerklein, J. Taeger, J. Hartig, J. Volkmann, R. Peach, D. Zeller

## Abstract

**Background:** Eye movement abnormalities are paramount in neurological disorders. However, unaided eye movement assessments lack granularity. Although videooculography (VOG) improves diagnostic accuracy, resource intensiveness preclude its broad use. To bridge this care gap, we here validate a framework for smartphone video-based nystagmography capitalizing on recent computer vision advances.

**Methods:** A recurrent convolutional network was fine-tuned for pupil tracking using >550 annotated frames: ConVNG. Slow phase velocity of optokinetic nystagmus was calculated in 10 subjects using both ConVNG and VOG. Equivalence of accuracy and precision was assessed using the “two one-sample t-test” (TOST) and Bayesian interval-null approaches.

**Results:** ConVNG tracking accuracy reached 9-15% of an average pupil diameter. SPV measurement accuracy was equivalent to VOG (p< .017; Bayes factors (BF) > 24). Average precision was 0.30° for ConVNG and 0.12° for VOG.

**Conclusions:** ConVNG enables smartphone video nystagmography with an accuracy comparable to VOG and precision approximately one order of magnitude higher than comparable ARKit applications. This serves as a blueprint for highly accessible tools with potential to accelerate progress toward precise and personalized Medicine.

## Background

Disturbances of ocular motility and coordination mark a highly interdisciplinary subject matter extending into the fields of neurology, ophthalmology and otology. Somewhat unique to motor physiology, eye movements can be grouped into a limited set of functional domains which specifically map onto distinct and/ or partly overlapping neural circuits^1^. These involve vestibular and visual afferents as well as neuronal networks encompassing the vast extent of cortex, brainstem and cerebellum. Therefore, most structural and functional pathologies of the peripheral and central nervous system manifest with eye movement disorders, lending to the notion of the eyes being a “window into the brain”.

Among those, *nystagmus (*from greek *“nystázein”*, to doze) has historically attracted the attention of clinicians and scientists due to its localizing and predictive value in various physiological and pathological contexts^2^. This is especially true in the context of vertigo and dizziness, which are among the most prevalent and economically challenging conditions world-wide^3,4^.

Over the last century, different approaches to register eye movements and in particular nystagmus have been developed, ranging from early eye photography to invasive (i. e. scleral search coils), electrical (e. g. electrooculography) and most recently, infrared and video based (i. e. videooculography (VOG) goggles) techniques (for a review see^1,5^). Besides documenting the mere presence and three-dimensional direction of nystagmus, the aforementioned methods enable to quantify slow phase velocity (SPV) as the quintessential descriptor of pathophysiology underlying nystagmus. Traditionally, SPV serves as the prime kinematic descriptor of nystagmus dynamics and is relevant to diagnostic^6,7^ and therapeutic decisions alike^8,9^. Common to VOG methods however is a high degree of resource intensiveness encompassing both monetary and educational aspects, hindering broad use in diverse clinical settings and especially outside of highly specialized laboratories, high-resource settings and academic infrastructure.

This can be viewed as a significant care gap, since recent clinical investigations convincingly demonstrate clinically relevant benefits of quantitative eye movement recordings for diagnosis^6,10–12^, prognostication^13^ and disease monitoring^9,14^. Moreover, yet unknown disease patterns could be identified within the granular kinematic feature space provided by e. g. videooculography, hinting at a largely untapped potential to derive performant physiomarkers to be included in clinical decision making^6,15^.

The episodic and often evanescent nature of many frequent neurological conditions presenting with eye movement abnormalities (e. g. vestibular migraine which affects up to 1% of the population^16^) is associated both with missed diagnoses and misdiagnosis^6,17^. Therefore, broadly available, point-of-care and longitudinal monitoring solutions to capture acute vertigo and improve diagnostic accuracy have received increasing attention^7,18^, with the idea of “telemetric nystagmography” dating back to 1991^19^. However, the aforementioned limitations of resource-intensiveness and lacking availability also apply to such devices, unfortunately minimizing their broad impact.

Recent shifts in health care landscapes necessitating remote, home use and telemedicine approaches^20^ are increasingly being answered with smartphone-based digital health applications shown to be capable of recording clinically meaningful eye and head movement data with acceptable accuracy and precision^21,22^ while performing on par with mobile eye-trackers for experimental applications^23^. Moreover, the rapidly evolving field of computer vision field has revolutionized the way in which behavior is measured using experimental^24,25^ as well as VOG recordings^26^ and clinical^27–29^ videos. Given the worldwide practiced tradition of clinical photo- and videography, especially in the field of Neurology and Ophthalmology, computer vision has the potential to unlock vast data treasures, likely containing digital biomarkers for prediction, prevention, prognostication and diagnostics^30^.

Undoubtedly, highly accessible, non-invasive, hardwareless and objective means to record health-related data in diverse contexts will promote personalized and precision medicine. Therefore, such developments are among the highest priorities in current digital health research portfolios^30,32,33^.

Here, we set out to develop a computationally inexpensive algorithmic framework based on the open-source computer vision tool DeepLabCut^24^ to extract SPV from smartphone nystagmus videos taken in realistic clinical scenarios and validate these measures against the current clinical gold standard, infrared VOG.

## Methods

See figure 1 for an illustration of the methodological workflow.

**Figure 1.**
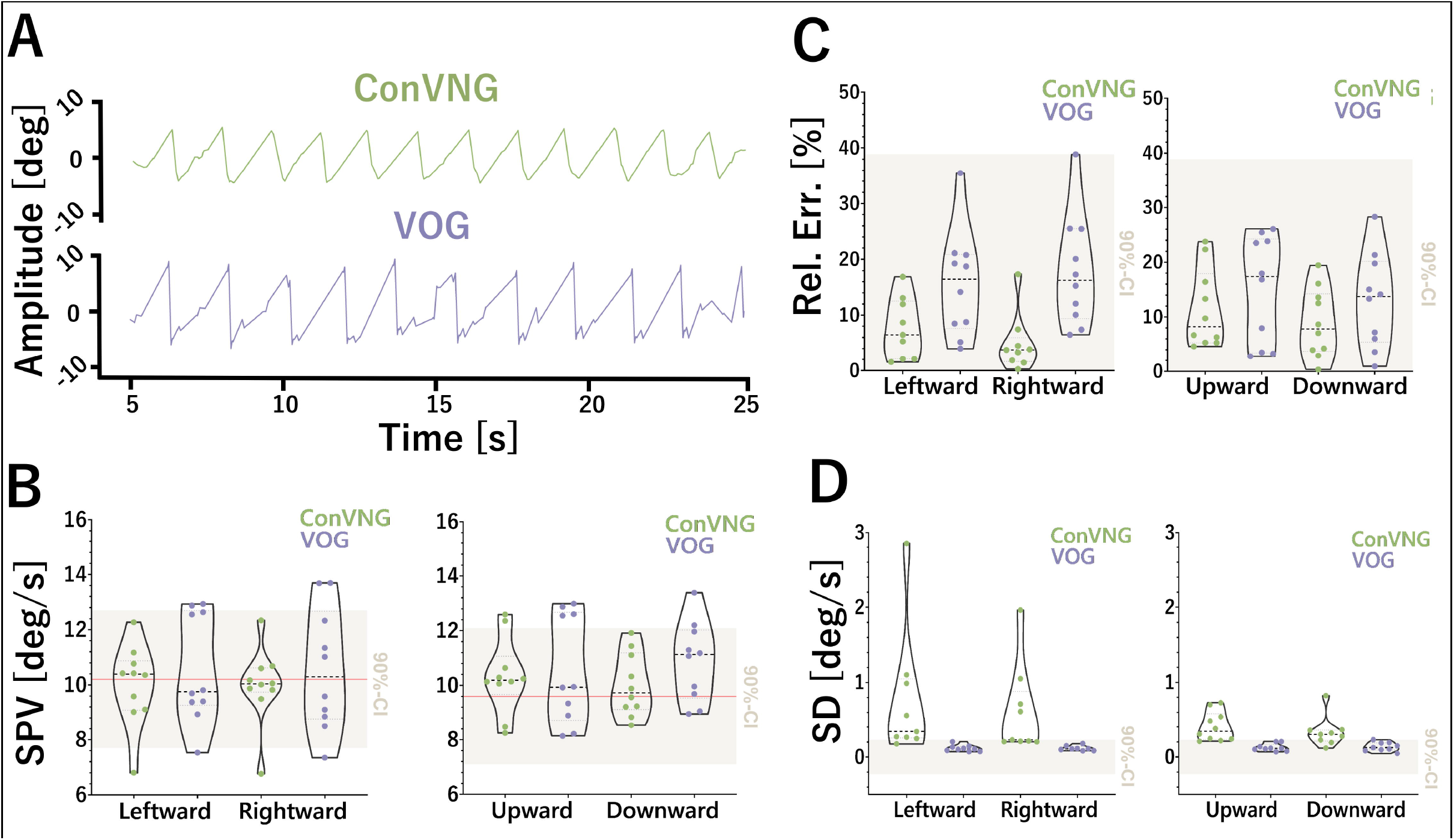
Workflow illustration. **A**. Illustration of standardized optokinetic stimulus presented on a smartphone screen. **B**. Exemplary frame of a tracked video, showing the typical camera perspective as well as ConVNG marker predictions, the color code of which is shown in **C**. L= left, R= right, P= pupil, L= lid, lat= lateral, 1-4 denote pupil marker position where 1 and 3 represent top and bottom (clock position 12 and 6) and 2 and 4 medial and lateral; e. g. “LP3” denotes left pupil bottom marker and “RLmid” the medial border of the right eyelids etc. **[not included in preprint version, please refer to the corresponding author for more information.] D**. Exemplary raw aggregate data plot and **E**., raw coordinate time-series plot from ConVNG-derived landmarks. Upper portion depicts vertical coordinates, lower portion horizontal coordinates. Note the already clearly recognizable “saw tooth” appearance typical for nystagmus in **E. F**. Median likelihood of predicted pupil labels in the out-of-sample validation approach using 10 independent videos. **G**. Exemplary likelihood plot from all landmarks derived of the same video as B. Except for “nosetop” marker, all landmarks are tracked with constantly high likelihoods (∼1.0).

### Tuning a recurrent convolutional neural network to perform pupil tracking

Utilizing the open-source framework DeepLabCut (DLC)^34^, a recurrent convolutional neural network (RCNN) based on a residual network architecture (ResNet 50) was trained to track a total of 17 landmarks delineating each pupil’s outlines at clock positions three, six, nine and twelve (n=8) as well as anatomically defined support points of interest in the face (eye lid borders, root of the nose, n=9, Fig. 1B). To train a maximally robust model, a total of 558 frames were extracted from >50 highly variable videos depicting ∼40 individuals (7-15 frames per individual). The videos were collected from the authors’ own or openly available video collections^35^ and showed eye movements and faces of diverse persons in various situations, lightings and camera settings, both in physiological and pathological contexts. To ensure broad coverage of possible pose patterns, the k-means algorithm implemented in DeepLabCut^24^ was selected for extraction of frames which were subsequently labelled by an expert annotator (MF). In order to exclude labelling errors which may negatively affect generalization, the labeled frames were plotted and quality-checked for accuracy and plausibility before the RCNN was trained on a 95% fraction of data leaving the remainder as a test set for later performance evaluation. No videos of the prospective cohort recruited to validate SPV measurements were included in this training dataset to ascertain clear separation between training, test and validation datasets throughout. The RCNN was fine-tuned using DLC’s default augmentation and ResNet 50 initialization weights. Sufficient convergence of the loss function was ensured with training iterations ranging between 350.000 to 750.000 in a total of 14 consecutive refinement iterations using unseen video material. Model performance was evaluated using a polypragmatic approach: first, by computing the mean Euclidean distance (MED) of user-annotated and RCNN-predicted labels^34^, second, by relating the sizes of MED and tracked region of interest as previously reported by our group^36^ and third, by systematically inspecting pupil marker likelihoods as a surrogate for the model’s prediction certainty in a fully independent (i. e. out-of-training) dataset of another 10 videos randomly sampled from an open-source, comprehensive eye movement disorder education library (courtesy Dan Gold)^35^.

### Eye movement recordings

#### A priori power analysis

Before subject recruitment, we carried out an a priori power calculation for two (paired) one-sided t-tests (TOST) powered to detect equivalence of SPV measurements. Assuming a β/α-ratio of 1.0 and a moderate effect size (d=0.65) due to the multivariate technical superiority of VOG over smartphone video (infrared vs. RGB sensor for contrast maximization, 220Hz vs. 30Hz temporal resolution, goggle/head vs. tripod mounted camera), at least 10 subjects were needed for a power >0.8. This cohort size matches well with comparable studies^22,31,37^.

#### Determination of intervals for equivalence testing

A smallest effect size of interest (SESOI)^38^ between 2-3°/s (i. e. 2.5°/s) was established in a consensus of the authors specializing in neurotological and neuroophthalmological conditions. This value was inferred so as to reflect a conservative estimate of a minimal SPV sufficient to generate perceptible oscillopsia in humans and to prompt further clinical work-up e. g. when encountered in screening for oculomotor and vestibular disorders, therefore having theoretical and practical relevance. Of note, SPVs higher than 5°/s are associated with a relevant interference with reading ability^39^, which is in our experience a rather liberal estimate. Also to improve statistical rigor, we decided to keep the more conservative value of 2.5°/s agreed upon in said consensus. Corresponding to approximately 25% of the estimated ground truth SPV, this value also matches with clinically used cut-offs to determine directional asymmetry of e. g. caloric nystagmus^40^. For relative error comparisons, a corridor of accepted measurement deviation was deducted from the deviation extremes measured with gold standard VOG after outlier testing in order to reduce the chance of misinterpreting an outlier as an acceptable deviation margin. These inferences were based on established methods to determine minimal meaningful effects in clinical studies^38,41^ and are in line with heuristic frameworks of SESOI definition^42,43^.

#### Subjects and ethics approval

For prospective validation, 10 healthy subjects (5 female) aged 25-44 years without significant neuroophthalmologic abnormalities, normal binocularity and an uncorrected visual acuity >0.6 were included. For the retrospective validation, cases with both VOG and frontoparallel eye movement video recording available from the same session were screened in the authors’ databases and collections. This query yielded two more subjects who had presented with downbeat nystagmus and congenital nystagmus to the lead author’s clinic between 2018 and 2019. Ethics approval was obtained from Julius-Maximilians University Wuerzburg’s ethics committee under the number 318/21.

#### Experimental setup

Experimental procedures were conducted in a naturally illuminated room at daytime. Subjects were seated in a distance of 40cm in front of a smartphone (iPhone XR, Apple, Cupertino) mounted on a standard tripod with the screen facing frontoparallely (Figure 1A). Screen orientation was switched 90° depending on the respective stimulus plane, i. e. using landscape mode for horizontal stimuli. Alignment of the screen with the horizontal and vertical axes was ascertained by using a built-in level of the tripod. The smartphone screen’s width was measured to be 14.0 cm, mapping onto 896 *points*, an abstract unit referencing the screen as a coordinate system used by Apple (https://developer.apple.com). This in turn results in a total angular viewing range of approximately 20°, a value which was chosen in accordance with standard VOG paradigms^44^. The height of the tripod was adjusted so as to align the screen’s center with the meridian of the subject’s eyes. The subjects were instructed to rest the back of their heads against a wall behind them or, in case of sitting freely, to hold the head as steady as possible while recording. The subjects’ interpupillary distance was measured with a distance ruler upon fixation of an object positioned to match the experimental approach (∼40cm) for later pixel-to-metric unit conversion.

#### Video recording

Eye movements were video recorded with smartphone cameras (iPhone Version 7 and XR Apple, Cupertino, CA, USA) at 1920×1080 pixels and a framerate of 30Hz, and in the case of one subject, 60Hz.

#### Videooculography

For gold standard monocular videooculography (VOG) recordings, the “EyeSeeCam Sci” (Version 8108847, EyeSeeTec, Munich, Germany^45,46^) was used. The camera’s spatiotemporal resolution is documented to be 188×120 px and 220 Hz, yielding a spatial resolution of 0.05 – 0.1° translating into an accuracy of approximately 1°. The VOG camera was connected to a Lenovo ThinkPad T470 Core i7 laptop running the proprietary OtoAccess version 1.5 recording software (Interacoustics, Middelfart, Denmark). The goggles’ headband was adjusted for a firm and snug fit. The camera was aligned in subject’s gaze straight ahead to center the reflection of the pupil in the image frame and subsequently calibrated for every individual using the built-in five-point laser grid projected onto a white wall located in one meter distance. Calibration plots were visually inspected by the experimenter (MF) before recordings were started in the software’s default “nystagmus” mode.

#### Stimulus material

To elicit nystagmus in a standardized way, we delivered a monochrome optokinetic nystagmus drum stimulus on the smartphone revolving at a constant rate of 555 points/ s (equalling 8.7cm/ s) along the screen, corresponding to an angular velocity of ∼12°/s (Figure 1A). This speed was chosen (i) on practical grounds as it allowed optimal stimulus pursuit^47,48^ as well as (ii) because it reflects the typical frequency of both physiologic caloric and pathological vestibular nystagmus encountered in daily practice^6,7,9,49^. Based on the assumption of a normal optokinetic response gain of 0.85 for horizontal and 0.80 for vertical stimuli in neurotypical subjects below the age of 50 years^48,50^, the ground truth of slow phase velocity was estimated to be 10.2°/s for horizontal and 9.6°/s for vertical directions.

#### Comparative nystagmus recordings

Since VOG goggles significantly obscured facial features essential for RCNN-based pupil tracking, nystagmus recordings were carried out in two back-to-back sessions taking place in a randomized order. To minimize intraindividual nystagmus heterogeneity between recording sessions, subjects were instructed to preferably follow fixation targets until they disappeared from the screen before refixation of a remote target on the opposite side of the screen. Nystagmus was recorded over 30 seconds in each of the four directions: left, right, up and down with breaks of two minutes between conditions for recuperation.

#### Deriving kinematic measurements from eye movement videos

##### Preprocessing

A striking inverse relationship of the RCNN’s tracking performance, as assessed by stability and plausibility of landmark predictions upon visual inspection of outputs, and spatial resolution of the videos (i. e. better tracking in lower resolutions) was observed. Evidently, 640×480px was the optimal resolution, retaining enough meaningful spatial information for landmark tracking while being associated with the most favorable tracking performance (Figure 1D, E, G). Therefore, all videos were downsampled to this resolution.

Kinematic analysis was implemented in Python using standard scientific analysis packages (pandas, sklearn, numpy, scipy). Due to the inexact alignment of video and stimulus onset, the DLC output was first filtered for times of interest (the first and last 5 seconds are cut from the analysis). Next, the data was cleaned using two steps: (1) we removed low likelihood marker data points (p<0.8, likelihood defined by DLC during prediction), (2) we removed impossible coordinates, i.e., where the markers for the pupil lie outside of the area spanned by the eye lid markers (rarely observed artefact). The tracked pupil marker positions (see diagram of markers in Figure 1B) were averaged for each eye to determine a pupil centroid, before using a bandpass filter (low cut = 0.5 Hz, high cut = 14 Hz), removing both high frequency noise and low frequency large scale movements (e. g. slow head or camera movements).

For transforming *pixels* into *metric units*, a conversion factor was derived per individual by dividing the actual interpupillary distance (IPD, average of three measurements from mid-pupil to mid-pupil) by the horizontal distance between both ConVNG tracked pupil centroids in pixels (average ∼60mm, in line with previous anthropometric data^51^), before conversion to degrees using angle= arctan (x/r), where r is the radius of the eye (defined as 12mm, in line with anthropometric data^52^). Using aforementioned IPD average instead of individual values did not significantly influence SPV calculations. The timeseries of each pupil (left and right, in degrees) is smoothed using a median filter (ndimage package, window length=3).

##### Classification of nystagmus direction

The direction of the nystagmus (horizontal or vertical) was determined by calculating a fast fourier transform of the x and y components of the pupil trajectory. If the absolute power is larger in the x-direction, then the nystagmus fast beating component is horizontal, and vice versa for vertical. Since the nystagmus may have a component in both the x-y directions (e. g. due to slightly oblique camera perspective), we computed the magnitude of the combined trajectories of x and y, 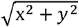. The resulting trajectories are further processed by performing a linear interpolation (interp, numpy) between the peaks and troughs (peak detection, scipy, prominence=1) of the nystagmus trajectory, producing a saw tooth signal.

##### Calculation of slow phase velocity

Slow phase velocity is the primary kinematic measure to characterize jerk nystagmus and therefore widely used for its diagnostic classification and monitoring alike^2,6,7,9^. Directional SPV comparisons are routinely used to determine relative asymmetries of caloric excitability of either labyrinth^40,53^. The slow phase velocity (SPV) for each eye is calculated by multiplying the median of the instantaneous gradient (gradient function, numpy) by the sampling frequency of the original video. The sign of the gradient (positive or negative) per plane indicates whether the slow phase velocity is leftward, rightward, upward or downward.

#### Statistical methods

Normality of datasets was examined using Kolmogorov-Smirnov testing and additional inspection of quartile (“Q-Q”-) plots so as to inform selection of appropriate display of data distributions as well as parametric or non-parametric analyses. For SPV analyses, binocular measurements resulting from ConVNG were averaged. For relative error calculations, both method’s absolute deviations from the plane-specific estimated ground truth were divided by the observed value. Equivalence was assessed using the two one-sample t-test (TOST) method^42^ with smallest effect sizes of interest^43^ derived as outlined earlier. Additionally, the Bayesian interval-null method for equivalence testing was used^54^ (see supplementary data). Outliers were removed using the robust regression and outlier removal (ROUT) method with a balanced coefficient Q= 1%^55^. For all statistical computations, GraphPad Prism Version 9, JASP Version 0.14.1^56^ and JAMOVI Version 2.2.5.0^57^, both with R plugins^58^ were used. Significance level was set at 5% (i. e. p< .05).

## Results

### An RCNN for tracking of pupils and facial landmarks

RCNN performance was evaluated in a tripartite approach. First, the mean Euclidean distances (MED) of user-annotated and RCNN-predicted labels on the training and test data subsets were measured. These were 2.22 and 6.12 pixels, a ratio reflecting acceptable to good generalization^34^. Second, an additional, pragmatic performance evaluation was carried out by relating the magnitude of the MED in the test set to the size of the tracked structure of interest as previously reported by our group^36^: given the average pupil diameters occupying 40-70 pixels throughout the used video material, MED was calculated to correspond to 9-15% of a pupil diameter and therefore confirmed to be acceptably small for further predictions (see supplementary videos 1 through 4 for exemplary labelled videos). Lastly, the median likelihood assigned to each of the eight pupil markers in a set of ten fully independent clinical eye movement videos randomly sampled from an open-source eye movement video collection^35^ was 0.85 (95% CI [0.82, 0.86], range: 0.2 – 0.96 before outlier detection and 0.55 – 0.96 after removal of four outlier values identified using ROUT method at a Q=1%, Fig 1F), demonstrating high out of sample model robustness. Upon further inspection, the lowest likelihood labels were yielded by one video. A lack of visual separability of pupil edge and an exceedingly dark iris could be identified as the most likely confounding factor.

### Accuracy and precision of slow phase velocity calculations

As the prime parameter to characterize intensity of jerk nystagmus, VOG-based SPV calculations are routinely used in clinical practice both in diagnostic^6,7,53^ and therapeutic^9,14,59^ contexts. To compare ConVNG’s aptitude to determine SPVs of experimentally standardized nystagmus, accuracy and precision were compared to the clinical gold standard method, infrared VOG.

#### Two one-sample T-Test

Within derived equivalence boundaries of ±2.5°/s, TOST revealed equivalence of SPV measures in all planes (leftward, upper limit T(9)=-4.5, p< .001, lower limit T(9)= 2.8, p= .01; rightward, upper limit T(9)= -4.0, p= .002, lower limit T(9)= 2.51, p= .017; upward, upper limit T(9)= -4.0, p= .002, lower limit T(9)= 3.34, p= .004; downward, upper limit T(9)= -7.5, p< .001, lower limit T(9)= 3.6, p= .003, Figure 2A-B).

**Figure 2.**
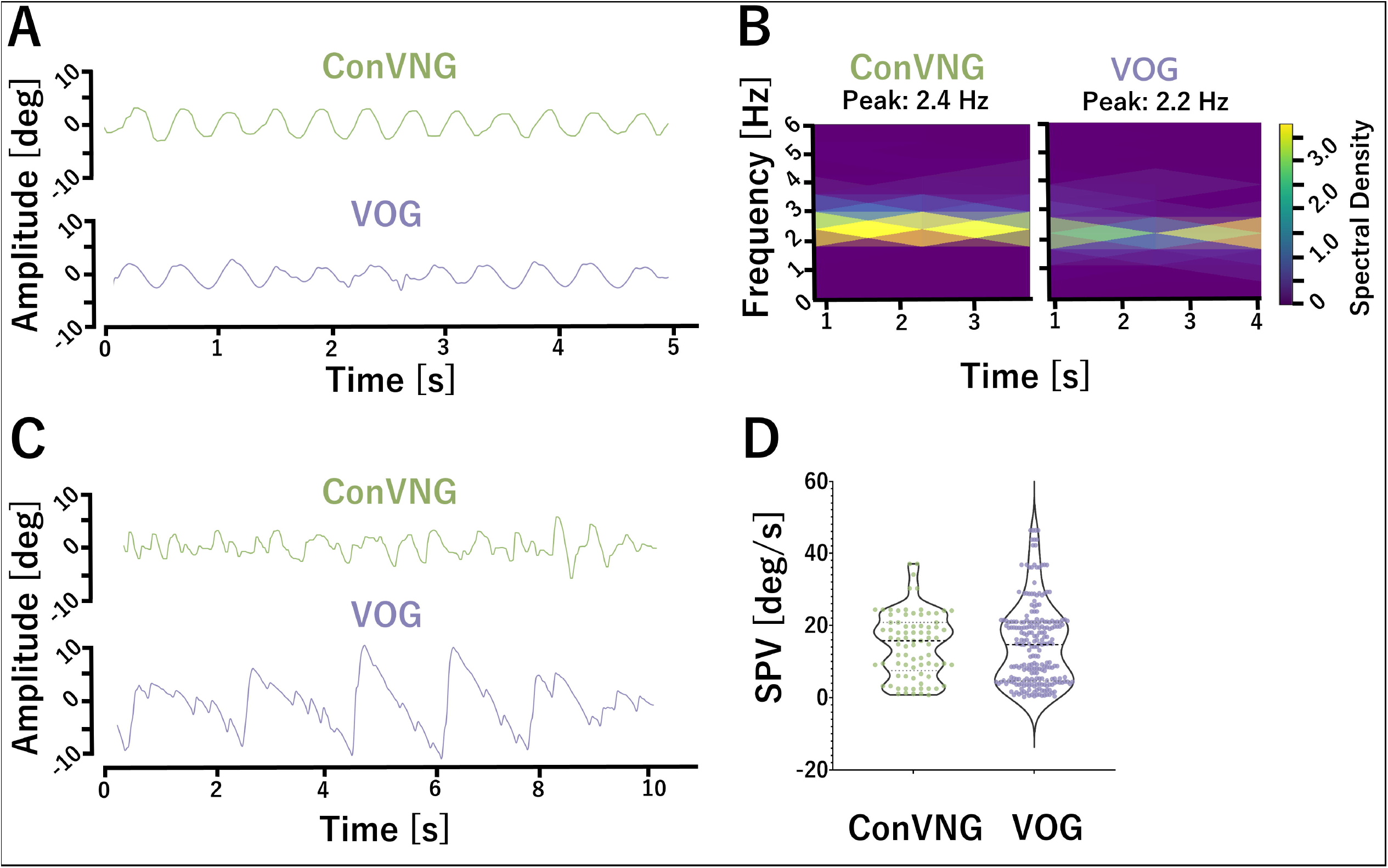
Validation. **A**. Exemplary traces derived from ConVNG and VOG for comparison. **B**. SPV values for both horizontal and vertical planes are shown in relation to the 90%-CI for equivalence testing. Red line denotes ground truth SPV value for horizontal (10.2°/s) and vertical (9.6°/s) planes. ConVNG values fall well within equivalence boundaries (±2.5°/s). **C**. Relative error values for accuracy calculation, in relation to the equivalence interval. ConVNG values fall well within equivalence boundaries (±38.9%). **D**. Standard deviations of sequential SPV measurements per plane for precision calculation, in relation to the equivalence interval. ConVNG values are significantly larger than the upper equivalence boundary, indicating lower precision (means: ConVNG 0.3°/s and VOG 0.12°/s).

To assess directional symmetry, which, besides absolute SPV values, is the principle readout for caloric nystagmus assessment in the clinical setting^40,53^, equivalence testing was also performed within methods and between stimulus directions. Per convention, 20-33% of directional asymmetry^40,53^ in the horizontal plane constitutes the cut-off for pathological findings (i. e. caloric paresis). In relation to the ground truth estimate of SPV, these percentages correspond to 2.0-3.4°/s, again rendering ±2.5°/s a reasonable equivalence interval. Conversely, the assumption of symmetry does not apply to vertical planes in various oculomotor domains^60,61^. Therefore, only horizontal symmetry was tested. TOST revealed equivalence of directional SPV measures in both methods (ConVNG, upper limit T(9)= -6.5, lower limit T(9)= 9.2; VOG, upper limit T(9)= -6.5, lower limit T(9)= 6.6, all p< .001).

The SPV errors per plane in relation to the estimated ground truth were leftward 11.8±14.4, rightward 9.5±15.3, upward 11.4±7.3, downward 8.9±6.3% (median 6.6±11.0%) for ConVNG and 15.6±9.5, 17.8±10, 15.1±9.8, 13.0±8.6% (median 15.1±9.2%) for VOG. For equivalence testing, the maximum deviation of goldstandard VOG (38.9%, survived ROUT at Q=1%) was used as an anchor point for 90%-CI assumption, revealing equivalence of methods in all comparisons (leftward, upper limit T(9)=-7.3, lower limit T(9)= 6.02; rightward, upper limit T(9)= -6.8, p= .001, lower limit T(9)= 4.4; upward, upper limit T(9)= -12.2, p< .001, lower limit T(9)= 10.1, p< .001; downward, upper limit T(9)= -14.9, lower limit T(9)= 12.0, all p< .001, Figure 1C).

To determine measurement precision, the medians of SPV standard deviations per plane were computed: 0.34, 0.23, 0.34 and 0.30°/s at a sampling rate of 30 Hz for ConVNG and 0.11, 0.11, 0.12 and 0.12°/s at 220 Hz for VOG (all after ROUT at Q=1%). Deriving an equivalence boundary of ±0.12°/s from the maximum standard deviation of goldstandard VOG, TOST yielded significant results against all lower bounds (all p<.02), while comparisons against the upper bounds were not significant (Figure 1D), meaning the data does not allow the conclusion of equivalence within the used interval.

Additionally, Bayesian approaches to equivalence testing were used, which are outlined in detail in supplementary data.

### Retrospective validation in an exemplary clinical cases

In order to further assess the validity of ConVNG, the authors revisited recent jerk nystagmus cases from their respective clinics with both video documentation and VOG data available from the same examination so as to construct a retrospective convenience sample for validation. Likely due to the broad availability of VOG in the authors’ clinical settings, rendering simultaneous video documentation optional, only two cases were identified fulfilling the search criteria. The first subject presented with a pendular nystagmus restricted to the horizontal plane. Since SPV measurement is not fully applicable in pendular/ sinusoidal nystagmus, frequency and amplitude relationships are used for evaluation, since their product ultimately influences visual acuity^1^. Since the nystagmus showed gaze-dependent shifts in intensity, selected signal portions with gaze straight ahead were used for comparison. Computed spectrograms revealed a peak frequency of 2.4Hz (ConVNG) and 2.2Hz (VOG), corresponding to an absolute deviation of 0.2Hz (relative deviation 9%, Figure 3A-B, supplementary video 5). The second subject showed a nystagmus purely beating in the vertical plane (downbeat nystagmus). Upward SPV measurements were 15.7±8.9°/s (ConVNG) versus 14.9±13.5°/s (VOG), constituting an absolute deviation of 0.8°/s (relative deviation 5%), a value falling well within the previously specified equivalence boundaries (Figure 3C-D, supplementary video 6).

**Figure 3.**
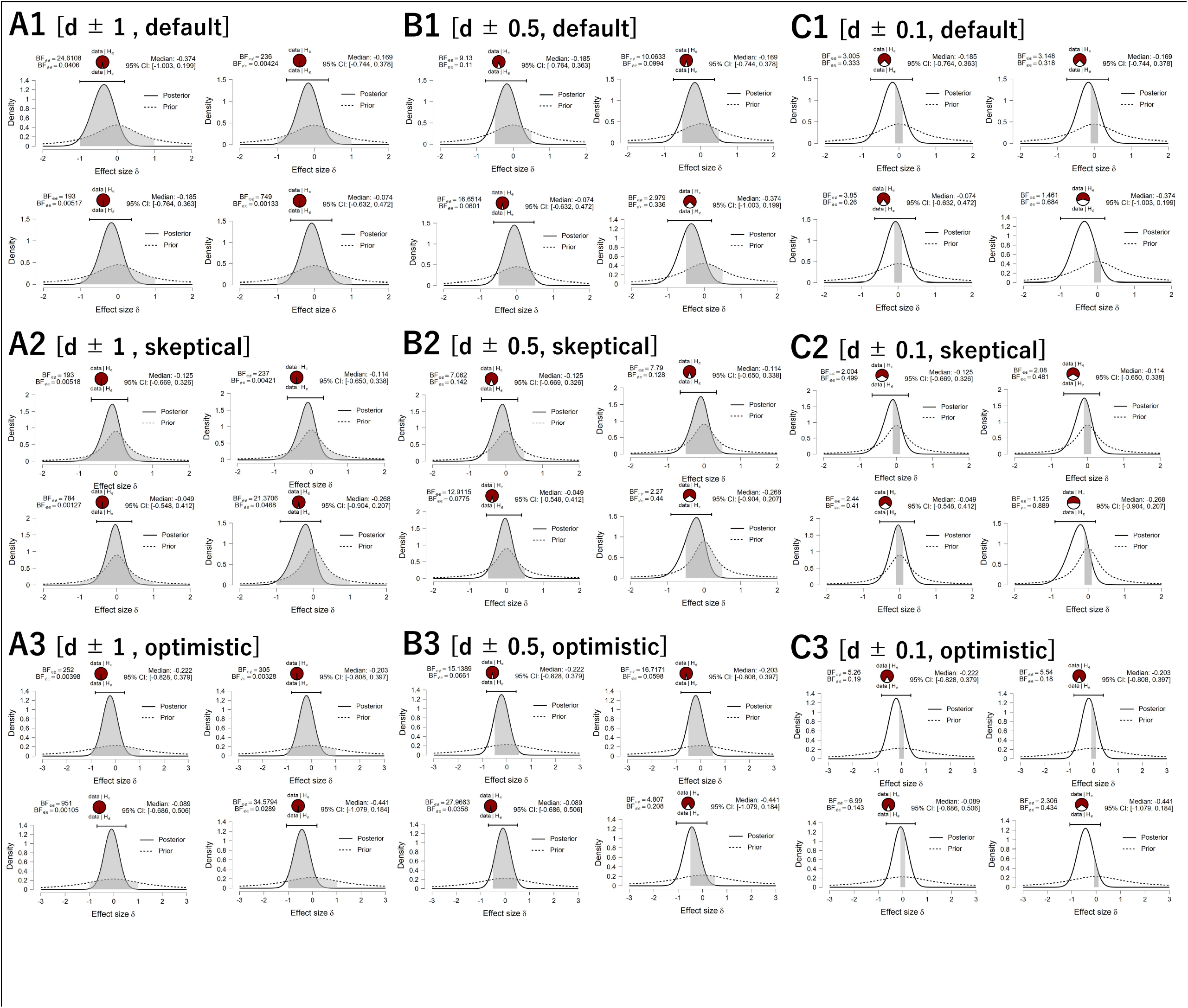
Retrospective validation. A. Traces from gaze straight ahead in case 1 and B., associated spectrograms. C. Traces from case 2 and D., associated SPV calculations.

## Discussion

Quantitative analysis of eye movements is a quintessential technique in the clinical toolbox with immediate diagnostic implications, especially in the complex and highly interdisciplinary context of oculomotor disorders and vertigo management^10,11^. Recent clinical investigations demonstrate that VOG-based assessments can double diagnostic certainty of vertigo syndromes in an emergency department setting^6^ and shorten time to diagnosis in episodic vertigo syndromes which usually do not coincide with patient presentations^7^. Furthermore, videonystagmography offers the unique capability of demasking subtle oculomotor deficits and can also double the diagnostic yield of oculomotor assessment in the setting of ataxia syndromes, thereby providing crucial diagnostic clues guiding targeted genetic diagnostics^12^. However, the resource intensiveness of current gold standard VOG precludes broad implementation, thereby demarcating factual and optimal management of neurological, otoneurological and neuroophthalmological patient populations.

To validate a highly accessible tool for video based nystagmus quantification, we chose a standardized, optokinetic instead of a caloric stimulus for two main reasons. Firstly, caloric nystagmus is inter- and intraindividually highly variable due to unforeseeable central habituation mechanisms on various timescales^40,53^, making a ground truth estimation impossible. This is in contrast to well established normative data available for optokinetic nystagmus^50^. Secondly, caloric nystagmus is significantly suppressed by visual fixation, necessitating either Frenzel goggles, which introduce spatial distortions in video recordings or dark conditions, which demand infrared cameras not routinely usable in most consumer grade cameras. Both factors have likely contributed to the only modest agreement between webcam video- and gold standard VOG-based nystagmus classification in a recent study by Reinhardt et al.^31^ In another closely related study focussing on a continuous monitoring aspect, Phillips and colleagues have demonstrated how novel, wearable technologies enable nystagmus detection and classification^18^. However, the extraction of quantitative measurements like SPV, a quintessential descriptor of nystagmus pathophysiology with established diagnostic^6,7^ and therapeutic^9,14^ implications, has not been successful in both studies^18,31^. In Young and colleagues’ study investigating a portable infrared VOG device as a vestibular event monitor, SPV measurements were not only acquired with high accuracy but could also differentiate common etiologies of episodic vertigo disorders^7^; however, a custom-built VOG device was needed.

The framework outlined here does not require any device except for a camera with at least 30Hz temporal and 640×480px spatiotemporal resolution, which is well below the current camera standard implemented widely in digital devices, webcams and the like. Making only few and pragmatic anthropomorphic assumptions regarding interpupillary distance and eyeball diameter^51,52,62^, computationally inexpensive standard algorithms like filtering, fast fourier transform, power spectral analysis, peak stripping and instantaneous gradient calculation (in line with standard SPV computations^63^) were used on the bilateral ConVNG-derived pupil centroid’s time series to identify nystagmus beating direction and calculate the average SPV per plane.

Despite a known bias of the TOST method towards non-equivalence decisions in small sample size scenarios like the one at hand^54^, it revealed equivalence of both methods’ SPV measurements, which converged with the outcomes of additional, explorative Bayesian testing.

Hence, these findings strongly support the notion that machine learning algorithms are not only capable of tracking pupils in laboratory acquired videos, but also of extracting time-series data accurate enough to enable meaningful and quantitative kinematic analyses in clinically relevant contexts. This was also true for the very small retrospective sample, which of course can only serve as a qualitative assessment. Still, in both experimentally uncontrolled instances (handheld camera recording, no tripod), ConVNG could be used to derive kinematic nystagmus descriptors with relative deviations in relation to VOG as little as 9% and 5%, respectively. Applying ConVNG to randomly sampled instances from a fully independent dataset^35^, yielded excellent out-of-sample robustness (median pupil label likelihood >0.85), underscoring feasibility of retrospective quantitative eye movement analysis regardless of experiment, setting or camera equipment.

This development adds to the growing interest in accessible, smart health care applications. As an important example, Parker and colleagues provided the proof-of-concept of an ARKit^37^-based iPhone application for video head-impulse testing^21,22^, thereby demonstrating feasibility of smartphone application-based quantitative eye and head movement recordings, albeit a considerable effort for manual postprocessing was needed. Using their ARKit-based application for gaze estimation, accuracy of 17% for horizontal and 27% for vertical planes (average 23%) and precision of 1.3° at lower gaze eccentricities was reported^22^. For comparison, precision of ConVNG reached 0.23-0.34°/s and, as expected, VOG reached precision as low as 0.11-0.12°/s. While equivalence of ConVNG and VOG in regards to precision could not be demonstrated, ConVNG’s absolute precision values of <0.34°/s are almost one order of magnitude smaller than ARKit’s documented gaze estimation^37^ and ∼75% smaller than the values achieved by Parker and colleagues^22^.

Notably, unlike VOG, both ARKit and ConVNG do not implement a formal calibration procedure. Referencing pupil position in both the video and orbital space is crucial for relating eye positions to stimulus or head positions, a procedure essential for gain value calculations (e. g. for video head impulse testing). Since SPV computation does not necessarily require absolute gaze positions, an implicit calibration procedure consisting of two anthropomorphic assumptions and a stable viewing distance of approximately 40cm as outlined above was sufficient to allow extraction of data with high accuracy. At the same time, the lack of calibration might partly explain why both ConVNG and ARKit’s^37^ precision metrics were clearly inferior to those of reference VOG. The possibility of calibration-free measurement with sufficient accuracy however is of particular relevance for at-home monitoring, especially in short-lasting, episodic conditions, which may practically not allow lengthy calibration procedures and associated user input-intensive device interaction.

Among the main methodological limitations of this study is the need for sequential eye movement recordings due to the fact that ConVNG is unable to track pupils and facial landmarks largely occluded or obscured by VOG goggles. However, in stark contrast to caloric nystagmus^31,53^, optokinetic nystagmus dynamics are significantly more stable between sequential measurements, making relevant intra-individual fluctuations confounding performance comparisons highly unlikely^1,48^. Additionally, we took precautions so as to avoid systematic biases by randomizing the order of experimental conditions for every individual. Still, this fact introduces an unforeseeable degree of biological variance, which needs to be considered when interpreting the equivalence testing and especially the comparisons of the retrospective comparisons. Another limitation of this study is the non-exhaustive representation of different nystagmus velocity conditions which would enable testing for correlations, linearity and systematical error distributions, but lie beyond the scope of this intended proof-of-concept. Instead, we defined ground truth values which closely match SPV values typically encountered in clinical practice to maximize validity^6,7^.

Relying on RGB videos, our approach is not expected to function in low light and low contrast settings, as was demonstrated in the out-of-sample validation. Combining the iPhone’s natural light and infrared sensors^21,37^, ARKit eye tracking promises to be more robust in these settings. However, in a series of experiments probing ARKit’s eye tracking capabilities (Taeger & Friedrich et al., unpublished data), we found that tracking performance significantly deteriorated in dim light conditions to the point of signal-to-noise ratios insufficient for SPV calculation. A recently proposed workflow based on convolutional neural networks applied to VOG-derived infrared eye videos has been associated with the most favorable pupil segmentation performance known to date^26^, however, as opposed to ConVNG it requires VOG equipment (high resolution infrared eye videos) in the first place. Albeit highly relevant for applications requiring visual fixation suppression, broadly accessible and available infrared camera technology remains an unmet need in clinical and experimental practice.

Taken together, our findings demonstrate that a 30Hz smartphone video can be sufficient for a specifically trained RCNN to extract quantitative eye movement parameters with an accuracy comparable to VOG and a precision higher than comparable ARKit implementations^22^. The presented pipeline’s computationally inexpensive and slender algorithmic architecture promotes broad deployment to both stationary and portable devices in diverse settings. Furthermore, ConVNG may stimulate large-scale retrospective investigations using medical video databases in unprecedented granularity.

Therefore, this approach holds tremendous potential for clinical and experimental applications ranging from telemetrics and at-home use to multivariate disease modeling for predictive, personalized and precision medicine. In our view, this will not supersede, but rather complement VOG in settings of low availability. To further validate the approach, larger-scale implementation efforts in even less controlled real-world settings, e. g. outpatient clinics and exploration of on-device analysis capabilities are warranted. To this end, we make the pretrained ConVNG model publicly available for the scientific community to build upon (https://doi.org/10.7910/DVN/GTUMAJ).

## Data Availability

Non-identifiable data and code sufficient to reproduce this study's main claims is available upon request to the corresponding authors. The RCNN model can be openly downloaded at the Harvard dataverse (doi:10.7910/DVN/GTUMAJ).

https://dataverse.harvard.edu/dataset.xhtml?persistentId=doi:10.7910/DVN/GTUMAJ

https://dataverse.harvard.edu/dataset.xhtml?persistentId=doi:10.7910/DVN/VG7XM2&version=DRAFT

## Supplementary data

### Explorative equivalence analysis using Bayes interval-null method

In light of its potential complimentary value in small sample and large effect size settings like the one at hand^54,64,65^, a more explorative approach to equivalence testing using Bayesian statistics was applied to this work’s key finding, i. e. SPV accuracy and precision. To this end, the smallest (i. e. most conservative) effect sizes, expressed as Cohen’s d values, corresponding to the raw interval boundaries (±2.5°/s) used for TOST were used to delineate a maximum equivalence interval around nil^65^. Systematic exploration of the equivalence likelihood was conducted in a bipartite approach: first, the maximum d values were multiplied by 0.5 and 0.1 so as to map out likelihoods of equivalence as a function of increasingly conservative interval boundaries. Second, due to insufficient data to inform a prior, a default Cauchy prior centered at 0 with a scale of 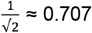 was used besides 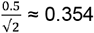 and 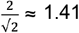 as more skeptical or “optimistic” variants, in line with previous comparative and explorative modeling in equivalence testing^54,66^.

In the Bayes interval-null method, the so called non-overlapping hypothesis (NOH) Bayes factor (BF) compares the likelihoods of the interval-null hypothesis (i. e. measurements are equivalent, H_0_) and the alternative hypothesis (i. e. effect sizes fall outside the equivalence region, suggesting non-equivalence, H_1_). NOH BF can be viewed as the degree, to which the data support values lying within versus outside of the equivalence intervals. The overlapping hypothesis (OH) BF compares the interval-null hypothesis against an unconstrained (i. e. no equivalence region around nil) alternative hypothesis and can be interpreted as the degree to which the data support the use of a specified equivalence region^65^.

Using an equivalence region of d ± 1.02 (smallest of all d-values corresponding to ±2.5°/s raw SPV interval boundaries used in TOST comparisons), OH BF for SPV comparisons per plane were 1.64, 1.64, 1.64 and 1.60, NOH Bayes factors 193.33, 235.65, 749.28 and 24.61, providing weak evidence for the overlapping hypothesis (i. e. restriction of parameter space to d ± 1.02) and extremely strong evidence in favor of equivalence. In other words, there is weak evidence in favor of the comparatively large equivalence boundary and given the data, the equivalence hypothesis is at least ∼25 times as likely as the non-equivalence hypothesis. At d ± 0.5, OH Bayes factors were 2.55, 2.64, 3.07, 1.40 and NOH Bayes factors were 3.0, 3.15, 3.9 and 1.46. At d ± 0.1, OH Bayes factors were 2.55, 2.64, 3.07 and 1.40, NOH Bayes factors were 3.0, 3.15, 3.85 and 1.46. Overall, these values provide moderate to strong evidence for an equivalence boundary of at least d ± 0.5, within which the equivalence hypothesis is ∼1.5- to 3.9-fold more likely than the alternative hypothesis. Neither adjusting priors to more skeptical 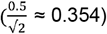 nor optimistic 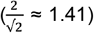 values substantially changes this conclusion (see supplementary figure 1 for systematic illustration).

For relative error equivalence testing, OH Bayes factors were 1.32 in all planes and NOH Bayes factors >142.285, providing weak evidence for the overlapping hypothesis and very strong evidence in favor of the equivalence hypothesis. At 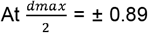, OH Bayes factors were 1.71, 1.71, 1.72 and 1.70 and NOH Bayes factors 34,67, 33.51, 45.21 and 25.1. 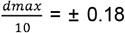, OH Bayes factors were 2.04, 2.02, 2.20 and 1.84, NOH Bayes factors were 2.53, 2.50, 2.83 and 2.20. Overall, this provides moderate to strong evidence in favor of the equivalence hypothesis within the smallest boundaries, d ± 0.18. As was the case for SPV measurements, adjusting priors to more skeptical 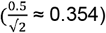 or optimistic 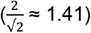 values yielded expected shifts of Bayes factors, however not substantially changing the conclusions drawn using the default prior (data not shown).

For precision estimates (d ± 0.14, corresponding to 0.12°/s minimal precision of VOG used in TOST), OH BF were 0.97, 1.04, 0.74 and 0.95, NOH BF were 0.97, 0.96, 0.72 and 0.95. At d ± 0.07 OH Bayes factors were 0.94, 1.0, 0.70, 0.92 and NOH BF were 0.93, 1.0, 0.69 and 0.91. At d ± 0.014 OH BF were 0.92, 0.99, 0.69, 0.90 and NOH BF were 0.92, 0.99, 0.69, 0.90. With NOH Bayes factors ranging between 0.69 and 1.0, consistent with weak evidence against the equivalence hypothesis with largely insufficient data to support a definitive conclusion.

Taken together, the Bayesian interval-null method’s outcomes across exploratory parametrizations of priors and equivalence region boundaries stand strongly in favor of the equivalence hypothesis in terms of clinically relevant accuracy intervals while providing largely inconclusive evidence for definitive conclusions for precision comparisons. Overall, these findings closely converge with TOST findings, thereby corroborating the equivalence hypothesis.

**Supplementary figure 1.**
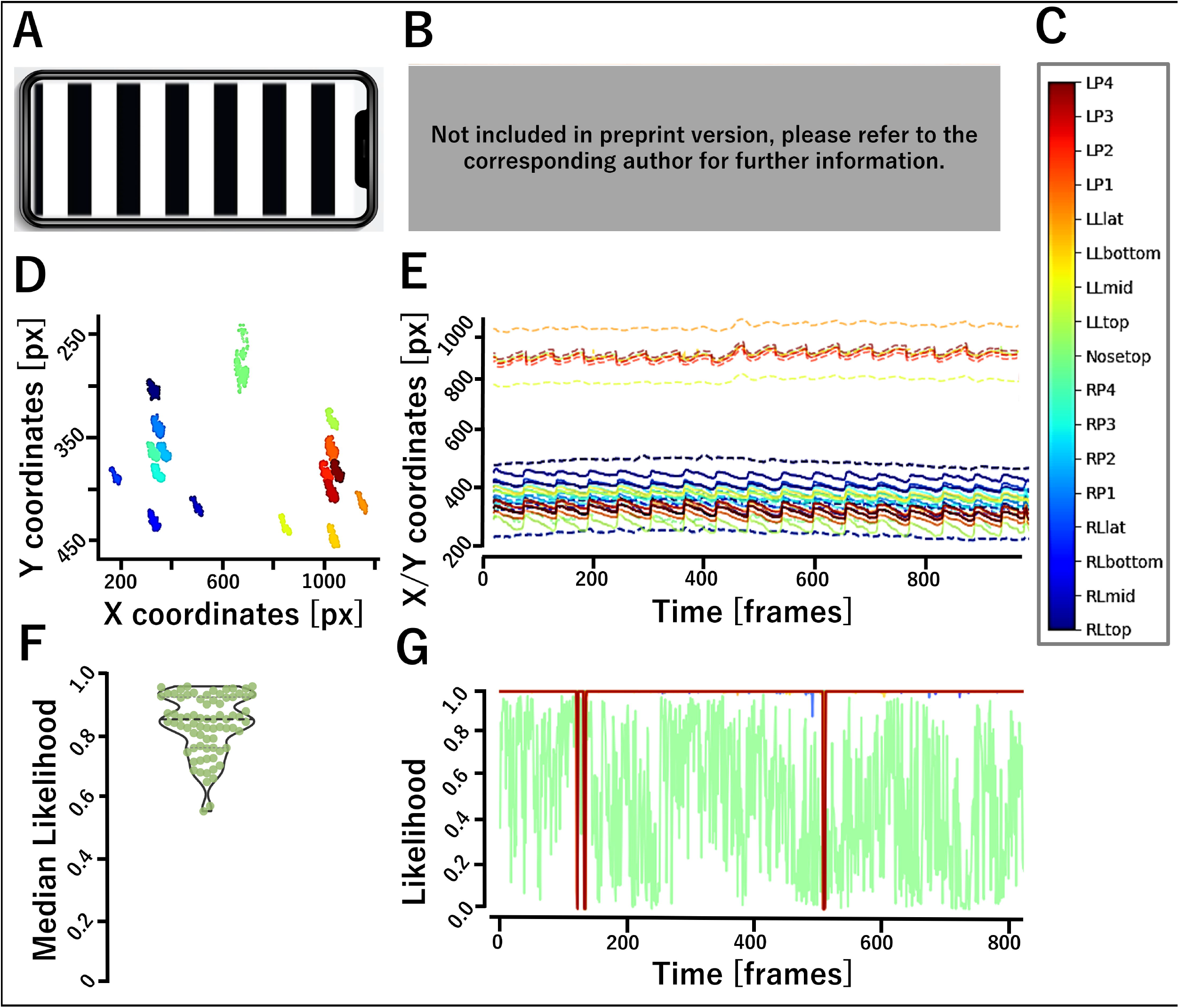
SPV equivalence testing using Bayes interval null method. Shown are the relationships of prior (dashed lines) and posterior (solid lines) likelihood distributions with equivalence regions shaded in grey. Rows (i. e. A-C) ordered by d-values for equivalence boundaries, columns (i. e. 1-3) ordered by prior estimates (see supplementary methods).

### Supplementary videos

**Supplementary Table 1.** Video descriptions. **[Not included in preprint version, please refer to the corresponding author for more information.]**

| ID | Content |
| --- | --- |
| 1-4 | Exemplary labeled videos of prospective cohort subjects showing nystagmus in all four planes. |
| 5 | Labeled video of Case 1, downbeat nystagmus |
| 6 | Labeled video of Case 2, pendular nystagmus |
| "OOSV"-Videos | Labeled videos randomly sampled from Dan Gold collection<sup>35</sup> used for out-of-sample validation. |

## Declarations

### Ethics and IRB approval, consent to participate and disclose

Subjects gave informed written consent which was conducted under the Julius-Maximilians University Wuerzburg’s ethics committee approval #318/21.

### Data and code availability

Non-identifiable data and code sufficient to reproduce this study’s main claims is available upon request to the corresponding authors. The RCNN model can be openly downloaded at the Harvard dataverse: https://dataverse.harvard.edu/dataset.xhtml?persistentId=doi:10.7910/DVN/GTUMAJ

### Authors’ contributions

MF established methods, obtained and analyzed subject and patient data, developed the models, conceptualized, drafted and revised the manuscript. JH, MB and JT obtained data and drafted the manuscript. RP analyzed subject and patient data and developed models and code. DZ supervised the experiment, obtained data and drafted the manuscript. ES analyzed subject and patient data drafted the manuscript. JV supervised the experiment, reviewed and revised the manuscript. All authors approved of the final version of the manuscript.

## Acknowledgements

The authors want to thank the subjects and patients involved in the study, Ann-Kathrin Karl, M. Sc., PD Dr. Andreas Zwergal, Egon Friedrich, Dr. Daniel R. Gold, MD and Dr. David Zee, MD for helpful remarks and fruitful discussions.

## Literature

1. Leigh, R. J. & Zee, D. S. The Neurology of Eye Movements. The Neurology of Eye Movements (Oxford University Press, 2015).

2. Tarnutzer, A. A. & Straumann, D. Nystagmus. Curr. Opin. Neurol. 31, 74–80 (2018).

3. Neuhauser, H. K. The epidemiology of dizziness and vertigo. Handb. Clin. Neurol. 137, 67–82 (2016).

4. Saber Tehrani, A. S. et al. Rising Annual Costs of Dizziness Presentations to U.S. Emergency Departments. Acad. Emerg. Med. 20, 689–696 (2013).

5. Wade, N. J. Pioneers of eye movement research. -Percept. 1, 33–68 (2010).

6. Nham, B. et al. Capturing vertigo in the emergency room: three tools to double the rate of diagnosis. J. Neurol. 269, 294–306 (2022).

7. Young, A. S. et al. Capturing acute vertigo: A vestibular event monitor. Neurology 92, e2743–e2753 (2019).

8. Luxon, L. Comparison of assessment of caloric nystagmus by observation of duration and by electronystagmographic measurement of slow-phase velocity. Br. J. Audiol. 29, 107–115 (1995).

9. Claassen, J. et al. Dalfampridine in patients with downbeat nystagmus--an observational study. J. Neurol. 260, 1992–1996 (2013).

10. Newman-Toker, D. E. et al. Quantitative video-oculography to help diagnose stroke in acute vertigo and dizziness: toward an ECG for the eyes. Stroke 44, 1158–1161 (2013).

11. Mantokoudis, G., Otero-Millan, J. & Gold, D. R. Current concepts in acute vestibular syndrome and video-oculography. Curr. Opin. Neurol. 35, 75–83 (2022).

12. Dupré, M., Hermann, R. & Froment Tilikete, C. Update on Cerebellar Ataxia with Neuropathy and Bilateral Vestibular Areflexia Syndrome (CANVAS). Cerebellum Lond. Engl. 20, 687–700 (2021).

13. Jung, I. & Kim, J.-S. Abnormal Eye Movements in Parkinsonism and Movement Disorders. J. Mov. Disord. 12, 1–13 (2019).

14. Abel, L. A., Walterfang, M., Stainer, M. J., Bowman, E. A. & Velakoulis, D. Longitudinal assessment of reflexive and volitional saccades in Niemann-Pick Type C disease during treatment with miglustat. Orphanet J. Rare Dis. 10, 160 (2015).

15. Halmagyi, G. M. et al. The Video Head Impulse Test. Front. Neurol. 8, 258 (2017).

16. Smyth, D., Britton, Z., Murdin, L., Arshad, Q. & Kaski, D. Vestibular migraine treatment: a comprehensive practical review. Brain awac264 (2022) doi:10.1093/brain/awac264.

17. Kerber, K. A. & Newman-Toker, D. E. Misdiagnosing the Dizzy Patient: Common Pitfalls in Clinical Practice. Neurol. Clin. 33, 565–viii (2015).

18. Phillips, J. S., Newman, J. L. & Cox, S. J. An investigation into the diagnostic accuracy, reliability, acceptability and safety of a novel device for Continuous Ambulatory Vestibular Assessment (CAVA). Sci. Rep. 9, 10452 (2019).

19. Wolf, S. R., Christ, P. & Haid, C. T. “Telemetric” Electronystagmography: A New Method for Examination of Nystagmus Outside the Clinic. Acta Otolaryngol. (Stockh.) 111, 374–381 (1991).

20. Chari, D. A., Wu, M. J., Crowson, M. G., Kozin, E. D. & Rauch, S. D. Telemedicine Algorithm for the Management of Dizzy Patients. Otolaryngol.--Head Neck Surg. Off. J. Am. Acad. Otolaryngol.-Head Neck Surg. 163, 857–859 (2020).

21. Parker, T. M. et al. Proof of Concept for an ‘eyePhone’ App to Measure Video Head Impulses. Digit. Biomark. 5, 1–8 (2021).

22. Parker, T. M. et al. Eye and Head Movement Recordings Using Smartphones for Telemedicine Applications: Measurements of Accuracy and Precision. Front. Neurol. 13, 789581 (2022).

23. Valliappan, N. et al. Accelerating eye movement research via accurate and affordable smartphone eye tracking. Nat. Commun. 11, 4553 (2020).

24. Mathis, A. et al. DeepLabCut: markerless pose estimation of user-defined body parts with deep learning. Nat. Neurosci. 21, 1281–1289 (2018).

25. Meyer, A. F., O’Keefe, J. & Poort, J. Two Distinct Types of Eye-Head Coupling in Freely Moving Mice. Curr. Biol. 30, 2116-2130.e6 (2020).

26. Yiu, Y.-H. et al. DeepVOG: Open-source pupil segmentation and gaze estimation in neuroscience using deep learning. J. Neurosci. Methods 324, 108307 (2019).

27. Rönn, A. J. et al. P 57 Markerless, video-based kinematic Tremor Analysis: machine vision in movement disorders. Clin. Neurophysiol. 137, e48 (2022).

28. Friedrich, M. et al. FV 23 POSe EstimatoR for Cervical Dystonia (POSER-CD): Automatized assessment of clinical severity and kinematic pathosignatures of Cervical Dystonia using convolutional neural networks. Clin. Neurophysiol. 137, e13 (2022).

29. Williams, S. et al. The discerning eye of computer vision: Can it measure Parkinson’s finger tap bradykinesia? J. Neurol. Sci. 416, 117003 (2020).

30. Esteva, A. et al. Deep learning-enabled medical computer vision. Npj Digit. Med. 4, 1–9 (2021).

31. Reinhardt, S., Schmidt, J., Leuschel, M., Schüle, C. & Schipper, J. VertiGo –a pilot project in nystagmus detection via webcam. Curr. Dir. Biomed. Eng. 6, (2020).

32. Taylor, K. I., Staunton, H., Lipsmeier, F., Nobbs, D. & Lindemann, M. Outcome measures based on digital health technology sensor data: data- and patient-centric approaches. Npj Digit. Med. 3, 1–8 (2020).

33. Esteva, A. et al. A guide to deep learning in healthcare. Nat. Med. 25, 24–29 (2019).

34. Nath, T. et al. Using DeepLabCut for 3D markerless pose estimation across species and behaviors. Nat. Protoc. 14, 2152–2176 (2019).

35. NOVEL - The Dan Gold Neuro-Ophthalmology Collection. https://novel.utah.edu/Gold/.

36. Knorr, S. et al. The evolution of dystonia-like movements in TOR1A rats after transient nerve injury is accompanied by dopaminergic dysregulation and abnormal oscillatory activity of a central motor network. Neurobiol. Dis. 154, 105337 (2021).

37. Greinacher, R. & Voigt-Antons, J.-N. Accuracy Assessment of ARKit 2 Based Gaze Estimation. in Human-Computer Interaction. Design and User Experience (ed. Kurosu, M.) 439–449 (Springer International Publishing, 2020). doi:10.1007/978-3-030-49059-1_32.

38. Anvari, F. & Lakens, D. Using anchor-based methods to determine the smallest effect size of interest. J. Exp. Soc. Psychol. 96, 104159 (2021).

39. Stahl, J. S., Plant, G. T. & Leigh, R. J. Medical treatment of nystagmus and its visual consequences. J. R. Soc. Med. 95, 235–237 (2002).

40. Gonçalves, D. U., Felipe, L. & Lima, T. M. A. Interpretation and use of caloric testing. Braz. J. Otorhinolaryngol. Engl. Ed. 74, 440–446 (2008).

41. Salas Apaza, J. A. et al. Minimal clinically important difference: The basics. Medwave 21, (2021).

42. Lakens, D. Equivalence Tests: A Practical Primer for t Tests, Correlations, and Meta-Analyses. Soc. Psychol. Personal. Sci. 8, 355–362 (2017).

43. Lakens, D., Scheel, A. M. & Isager, P. M. Equivalence Testing for Psychological Research: A Tutorial. Adv. Methods Pract. Psychol. Sci. 1, 259–269 (2018).

44. Bertolini, G. et al. Gaze Holding in Healthy Subjects. PLoS ONE 8, e61389 (2013).

45. Bartl, K., Lehnen, N., Kohlbecher, S. & Schneider, E. Head impulse testing using video-oculography. Ann. N. Y. Acad. Sci. 1164, 331–333 (2009).

46. Schneider, E. et al. EyeSeeCam: an eye movement-driven head camera for the examination of natural visual exploration. Ann. N. Y. Acad. Sci. 1164, 461–467 (2009).

47. Seferlis, F. et al. Age related changes in ocular motor testing in healthy subjects. J. Vestib. Res. Equilib. Orientat. 25, 57–66 (2015).

48. Valmaggia, C. et al. Age related change of optokinetic nystagmus in healthy subjects: a study from infancy to senescence. Br. J. Ophthalmol. 88, 1577–1581 (2004).

49. Pietkiewicz, P., Pepaś, R., Sułkowski, W. J., Zielińska-Bliźniewska, H. & Olszewski, J. Electronystagmography versus videonystagmography in diagnosis of vertigo. Int. J. Occup. Med. Environ. Health 25, 59–65 (2012).

50. Kullmann, A. et al. Normative data for ages 18-45 for ocular motor and vestibular testing using eye tracking. Laryngoscope Investig. Otolaryngol. 6, 1116–1127 (2021).

51. Pointer, J. S. The interpupillary distance in adult Caucasian subjects, with reference to ‘readymade’ reading spectacle centration. Ophthalmic Physiol. Opt. J. Br. Coll. Ophthalmic Opt. Optom. 32, 324–331 (2012).

52. Bekerman, I., Gottlieb, P. & Vaiman, M. Variations in eyeball diameters of the healthy adults. J. Ophthalmol. 2014, 503645 (2014).

53. Shepard, N. T. & Jacobson, G. P. The caloric irrigation test. Handb. Clin. Neurol. 137, 119–131 (2016).

54. Linde, M., Tendeiro, J. N., Selker, R., Wagenmakers, E.-J. & van Ravenzwaaij, D. Decisions about equivalence: A comparison of TOST, HDI-ROPE, and the Bayes factor. Psychol. Methods (2021) doi:10.1037/met0000402.

55. Motulsky, H. J. & Brown, R. E. Detecting outliers when fitting data with nonlinear regression –a new method based on robust nonlinear regression and the false discovery rate. BMC Bioinformatics 7, 123 (2006).

56. JASP Team. JASP. (2022).

57. The jamovi project (2021). jamovi (Version 1.6) [Computer Software]. Retrieved from https://www.jamovi.org.

58. R Core Team. R: A language and environment for statistical computing. (2021).

59. Claassen, J. et al. A randomised double-blind, cross-over trial of 4-aminopyridine for downbeat nystagmus--effects on slowphase eye velocity, postural stability, locomotion and symptoms. J. Neurol. Neurosurg. Psychiatry 84, 1392–1399 (2013).

60. Pierrot-Deseilligny, C. & Milea, D. Vertical nystagmus: clinical facts and hypotheses. Brain J. Neurol. 128, 1237–1246 (2005).

61. Murasugi, C. M. & Howard, I. P. Up-down asymmetry in human vertical optokinetic nystagmus and afternystagmus: contributions of the central and peripheral retinae. Exp. Brain Res. 77, 183–192 (1989).

62. MacLachlan, C. & Howland, H. C. Normal values and standard deviations for pupil diameter and interpupillary distance in subjects aged 1 month to 19 years. Ophthalmic Physiol. Opt. J. Br. Coll. Ophthalmic Opt. Optom. 22, 175–182 (2002).

63. Torok, N. & Derbyshire, A. J. Computation of the Nystagmogram. Acta Otolaryngol. (Stockh.) 65, 70–78 (1968).

64. van Ravenzwaaij, D., Monden, R., Tendeiro, J. N. & Ioannidis, J. P. A. Bayes factors for superiority, non-inferiority, and equivalence designs. BMC Med. Res. Methodol. 19, 71 (2019).

65. Lakens, D., McLatchie, N., Isager, P. M., Scheel, A. M. & Dienes, Z. Improving Inferences About Null Effects With Bayes Factors and Equivalence Tests. J. Gerontol. Ser. B 75, 45–57 (2020).

66. Fornacon-Wood, I. et al. Understanding the Differences Between Bayesian and Frequentist Statistics. Int. J. Radiat. Oncol. Biol. Phys. 112, 1076–1082 (2022).

